# An ecological examination of early adolescent e-cigarette use: A machine learning approach to understanding a health epidemic

**DOI:** 10.1101/2023.06.16.23291513

**Authors:** Alejandro L. Vázquez, Cynthia M. Navarro Flores, Byron H. Garcia, Tyson S. Barrett, Melanie M. Domenech Rodríguez

## Abstract

E-cigarette use among adolescents is a national health epidemic spreading faster than researchers can amass evidence for risk and protective factors and long-term consequences associated with use. New technologies, such as machine learning, may assist prevention programs in identifying at-risk youth and potential targets for intervention before adolescents enter developmental periods where e-cigarette use escalates. The current study utilized machine learning algorithms to explore a wide array of individual and socioecological variables in relation to patterns of lifetime e-cigarette use during early adolescence (i.e., exclusive, or with tobacco). Extant data was used from 14,346 students middle school students (M_age_ = 12.5, SD = 1.1; 6^th^ and 8th grades) who participated in the Utah Prevention Needs Assessment survey. Students self-reported their substance use behaviors and related risk and protective factors. Machine learning algorithms examined 112 individual and socioecological factors as potential classifiers of lifetime e-cigarette use outcomes. The elastic net algorithm achieved outstanding classification for lifetime exclusive (AUC = .926) and dual use (AUC = .944) on a validation test set. Six high value classifiers were identified that varied in importance by outcome: Lifetime alcohol or marijuana use, perception of e-cigarette availability and risk, school suspension(s), and perceived risk of smoking marijuana regularly. Specific classifiers were important for lifetime exclusive (parent attitudes regarding student vaping, best friend[s] tried alcohol or marijuana) and dual use (best friend[s] smoked cigarettes, lifetime inhalant use). Our findings provide specific targets for the adaptation of existing substance use prevention programs to address early adolescent e-cigarette use.

## Introduction

E-cigarette use among adolescents is a national epidemic [1]. The popularity of these devices has spread faster than health researchers could amass evidence for the potential deleterious effects of e-cigarette use. As the prevalence of traditional cigarette smoking among U.S. adolescents has declined, e-cigarette use, or vaping, has become the most commonly used form of nicotine uptake among youth in the U.S. [2]. Adolescents’ decisions to engage in e-cigarette use may be understood through an ecological framework that accounts for complex interactions between spheres of influence [3]. Research is underway to identify individual and socioecological risk-factors associated with e-cigarette use [4–11]. However, this literature has prominently focused on high school samples resulting in a dearth of knowledge regarding e-cigarette risk-factors during early adolescence. Identifying factors associated with the emergence of e-cigarette use during early adolescence may facilitate intervention prior to developmental periods where use escalates (i.e., middle to late adolescence [12]). These efforts may be bolstered by new methodologies that allow researchers to efficiently explore the importance of a wide range of variables in relation to e-cigarette use [13]. In this study we use machine learning algorithms to simultaneously consider a large number of individual and socioecological factors in relation to patterns of e-cigarette usage among middle school students [7].

The use of the e-cigarettes has been touted as a healthier alternative to tobacco cigarettes, despite their delivery of nicotine and other potentially harmful chemicals [14]. A major concern of nicotine consumption during early adolescence is the possible alteration of function in the brain’s reward systems at a sensitive developmental period, in ways that can increase risk for other substance use, mood disorders, and difficulties with concentration and learning [14]. In addition to nicotine-related risks, other carcinogenic agents found in chemicals in the e-liquid as well as those produced in the vaporizing product or even associated with the e-cigarette materials (i.e., nickel, chromium, cadmium; [14]). Chemicals used to flavor e-liquid have also been found to have sufficiently high toxicity to warrant medical concerns [15] or even cause death [16]. The possible harms of e-cigarettes go well beyond exposure to nicotine.

Researchers have documented complex relationships between individual (e.g., academic performance, substance use, perceptions of use) and socioecological (e.g., access, advertisements, peer and parental factors) influences implicated in e-cigarette use during middle to late adolescence [7–9]. Less is known about e-cigarette use risk-factors during early adolescence. The early adolescent e-cigarette literature has predominately focused on the prevalence and reasons for use, or factors associated with susceptibility rather than initiation [12,17–20]. Studies examining adolescent e-cigarette use have also had a narrow focus when considering potential individual and socioecological influences on adolescent e-cigarette use. For example, studies commonly focus on specific adolescent attitudes (e.g., perceived danger of e-cigarettes and tobacco), substance use behaviors (e.g., alcohol, tobacco, marijuana), aspects of the environment (e.g., access, advertisements), and social influences (e.g., peer and parental e-cigarette or cigarette use [7–9,12,19]). Research is needed to examine these risk-factors in conjunction with a broader array of influences traditionally associated with early substance use (e.g., anti-social behavior; parenting practices; school involvement, performance, environment; community attachment, norms, drug use, delinquency; [21–25]).

It is also important to consider patterns of use when identifying correlates of early adolescent vaping. Research suggests risk-factors vary between youth who have utilized e-cigarettes exclusively and those who have used them in combination with tobacco, with dual use being associated with greater behavioral problems (i.e., lifetime use; *M* = 14.6 years, *SD* = 0.7) and substance use (i.e., lifetime alcohol, marijuana, drug use prescription drug misuse; 9^th^ and 12^th^ graders; [7,26]). Exclusive use may represent adolescent using e-cigarettes as a “safer” alternative to traditional tobacco cigarettes [19]. Dual use may be associated with tobacco cessation or recreational use in conjunction with other substances [19,26]. Research has yet to determine whether differences in risk-factors for exclusive or dual e-cigarette use exist during early adolescence.

Methodological challenges may explain the limited number of studies examining a broad array of correlates of e-cigarette use during early adolescence. For example, lifetime e-cigarette is a low base rate behavior during early adolescence relative to later developmental periods [26]. Prevalence rates are even lower when researchers examine exclusive and dual e-cigarette use relative to general lifetime use [7]. Furthermore, limitations associated with traditional statistical methodologies may pose a barrier to examining the broad array of potential factors implicated in early adolescent e-cigarette use (e.g., statistical power issues, multicellularity; familywise error rate[13]). These limitations can be addressed with large datasets, however meeting statistical assumptions for multicollinearity and reducing family-wise error may limit the number of potential risk-factors that can be simultaneously considered in relation to early adolescent e-cigarette use.

Machine learning may facilitate the examination of factors associated with early adolescent e-cigarette use. Machine learning provides an efficient method of simultaneously examining large numbers of variables representing youth individual and socioecological factors to determine their *importance* in classifying substance use [13]. Within the context of machine learning, variable importance refers to the relative ability for variables to reduce the error in models’ predictions of group membership (e.g., exclusive e-cigarette user or non-user) compared to other covariates in the model [27]. Elastic net, random forest, k-nearest neighbors, and neural networks are examples of common algorithms that are capable of provided suprior accuracy in classifying lifetime substance use relative to traditional logistic regression [13,28]. Each of these algorithms approach classification with contrasting linear (i.e., elastic net) and nonlinear (i.e., random forest, k-nearest neighbors, neural networks) methods, providing the opportunity to identify the algorithm that best performs the classification task for each outcome [27].

Identifying high value correlates of e-cigarette initiation during early adolescence could improving our ability to identify at-risk youth prior to developmental periods were the prevalence and frequency of vaping escalates (i.e., middle to late adolescence; [12]). While machine learning may provide an additional tool for informing substance use prevention effort [29], few studies have utilized machine learning to identify factors associated with patterns of early e-cigarette use (i.e., lifetime exclusive use, dual use with tobacco) or determined which method provides the best classification accuracy. Prior applications of machine learning have predominately focused on unstructured data (i.e., pictures, text) to classify e-cigarette use [30,31]. While a recent study trained machine learning algorithms on survey data collected on older teens (M_age_ =15.36 years old; *SD* = 1.85), this research had a narrow focus on tobacco related substance use predictors that precludes a broader understanding of factors associated with early vaping initiation (i.e., LASSO and Random Forest; [32]). Thus, our aim was to (a) explore a wide array of factors using machine learning to identify important classifiers of lifetime exclusive e-cigarette and dual use within a sample of middle school students, (b) and identify the algorithm that best performs the classification task. These analyses can help quantify the relative importance of predictors and established the extent to which e-cigarette use can be classified by individual and socioecological factors.

## Materials and method

The current study utilized data from the Utah Student Health and Risk Prevention (SHARP) survey project, which has been collecting and disseminating information on substance use prevalence and related behaviors since 2007 (Utah Department of Human Services [UDHS]; [33]). SHARP was developed as a collaboration between multiple state agencies with the purpose of assessing risk and protective factors for problem behaviors among Utah middle and high school students. Students complete the Utah Prevention Needs Assessment (PNA) survey biannually, during the spring of odd numbered years, as a part of the SHARP survey project. The PNA survey gathers statewide data on substance use and individual/socioecological factors that influence the use of alcohol, tobacco, and other drugs. PNA surveys are implemented and used to inform statewide prevention policy and programming across the United States. Surveys are completed in schools and are self-administered using paper and pencil. The present study used data collected during the Spring of 2017 (i.e., March-June 2017) as part of a PNA survey in Utah. Parents provided written consent for their child to participate in the survey. Parents of youths that did not consent to their child participating in the PNA were not administered the survey. Student also provided verbal assent prior to participating in the PNA survey. Participation in the survey was voluntary and students could opt to participate in an alternative activity or discontinue at any time. The Utah State University Institutional Review Board approved secondary analyses of the 2017 Utah PNA survey data as non-human subjects research as participants could not be re-identified (protocol #10108). Previous research has utilized similar statewide school-based samples to identify factors associated with e-cigarette use among adolescents across the U.S. (e.g., Hawaii, Texas, Connecticut, New Jersey; [6–9,20,34]).

The current study focused on 14,346 middle school students (i.e., 6^th^ and 8th grade) that participated in the 2017 Utah PNA survey. Participants were approximately 12 years old on average (*M* = 12.5; *SD* = 1.1), were relatively balanced on sex (girls; n = 7,532, 52.5%) and 6^th^ grade (n = 7,473, 52.1%), and were predominantly White (9,491, 71%). Nearly a third of students attended school within Salt Lake County (*n* = 4,173, 29.1%) in Utah. Youths in this sample reported a 9.4% (*n* = 1,343) prevalence of lifetime e-cigarette use and 5.4% (*n* = 784) tobacco use. Students largely reported abstaining from both tobacco and e-cigarette use (91%; *n* = 13,003) and reported greater lifetime exclusive e-cigarette use (5.5%; *n* = 791) relative to exclusive tobacco use (1.6%; *n* = 232). Within the sample, 3.8% (*n* = 552) of students reported dual lifetime use of tobacco and e-cigarettes. See Table 1 for sample demographic information by outcomes.

**Table 1.**
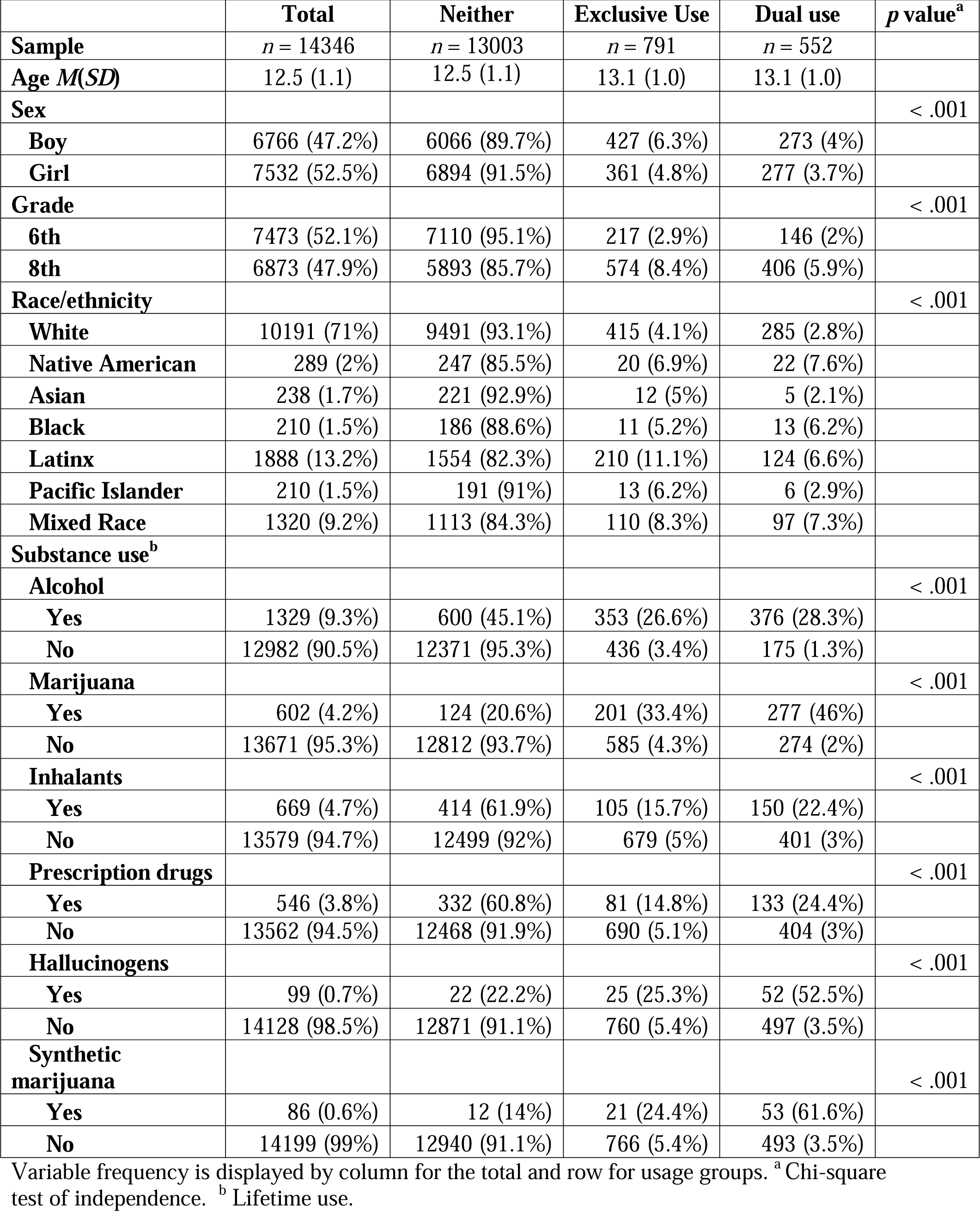
Student Demographics by Lifetime E-Cigarette Use Groups.

| **Table 1. Student Demographics by Lifetime E-Cigarette Use Groups.** |  |  |  |  |  |
| --- | --- | --- | --- | --- | --- |
|  | **Total** | **Neither** | **Exclusive Use** | **Dual use** | ***p* value<sup>a</sup>** |
| **Sample** | *n* = 14346 | *n* = 13003 | *n* = 791 | *n* = 552 |  |
| **Age *M*(*SD*)** | 12.5 (1.1) | 12.5 (1.1) | 13.1 (1.0) | 13.1 (1.0) |  |
| **Sex** |  |  |  |  | < .001 |
| **Boy** | 6766 (47.2%) | 6066 (89.7%) | 427 (6.3%) | 273 (4%) |  |
| **Girl** | 7532 (52.5%) | 6894 (91.5%) | 361 (4.8%) | 277 (3.7%) |  |
| **Grade** |  |  |  |  | < .001 |
| **6th** | 7473 (52.1%) | 7110 (95.1%) | 217 (2.9%) | 146 (2%) |  |
| **8th** | 6873 (47.9%) | 5893 (85.7%) | 574 (8.4%) | 406 (5.9%) |  |
| **Race/ethnicity** |  |  |  |  | < .001 |
| **White** | 10191 (71%) | 9491 (93.1%) | 415 (4.1%) | 285 (2.8%) |  |
| **Native American** | 289 (2%) | 247 (85.5%) | 20 (6.9%) | 22 (7.6%) |  |
| **Asian** | 238 (1.7%) | 221 (92.9%) | 12 (5%) | 5 (2.1%) |  |
| **Black** | 210 (1.5%) | 186 (88.6%) | 11 (5.2%) | 13 (6.2%) |  |
| **Latinx** | 1888 (13.2%) | 1554 (82.3%) | 210 (11.1%) | 124 (6.6%) |  |
| **Pacific Islander** | 210 (1.5%) | 191 (91%) | 13 (6.2%) | 6 (2.9%) |  |
| **Mixed Race** | 1320 (9.2%) | 1113 (84.3%) | 110 (8.3%) | 97 (7.3%) |  |
| **Substance use<sup>b</sup>** |  |  |  |  |  |
| **Alcohol** |  |  |  |  | < .001 |
| **Yes** | 1329 (9.3%) | 600 (45.1%) | 353 (26.6%) | 376 (28.3%) |  |
| **No** | 12982 (90.5%) | 12371 (95.3%) | 436 (3.4%) | 175 (1.3%) |  |
| **Marijuana** |  |  |  |  | < .001 |
| **Yes** | 602 (4.2%) | 124 (20.6%) | 201 (33.4%) | 277 (46%) |  |
| **No** | 13671 (95.3%) | 12812 (93.7%) | 585 (4.3%) | 274 (2%) |  |
| **Inhalants** |  |  |  |  | < .001 |
| **Yes** | 669 (4.7%) | 414 (61.9%) | 105 (15.7%) | 150 (22.4%) |  |
| **No** | 13579 (94.7%) | 12499 (92%) | 679 (5%) | 401 (3%) |  |
| **Prescription drugs** |  |  |  |  | < .001 |
| **Yes** | 546 (3.8%) | 332 (60.8%) | 81 (14.8%) | 133 (24.4%) |  |
| **No** | 13562 (94.5%) | 12468 (91.9%) | 690 (5.1%) | 404 (3%) |  |
| **Hallucinogens** |  |  |  |  | < .001 |
| **Yes** | 99 (0.7%) | 22 (22.2%) | 25 (25.3%) | 52 (52.5%) |  |
| **No** | 14128 (98.5%) | 12871 (91.1%) | 760 (5.4%) | 497 (3.5%) |  |
| **Synthetic marijuana** |  |  |  |  | < .001 |
| **Yes** | 86 (0.6%) | 12 (14%) | 21 (24.4%) | 53 (61.6%) |  |
| **No** | 14199 (99%) | 12940 (91.1%) | 766 (5.4%) | 493 (3.5%) |  |

Variable frequency is displayed by column for the total and row for usage groups. <sup>a</sup> Chi-square test of independence. <sup>b</sup> Lifetime use.

### Measures

#### Individual and socioecological variables

The measures utilized in the current study have been traditionally used and reported by the SHARP survey project as individual items [33]. Variables examined in the current study have been identified as being theoretically and/or empirically important factors in the substance use literature. We decided to examine individual items to provide a nuanced understanding of e-cigarette use risk-factors [13]. Variables included a wide array of factors representing *individual* (i.e., antisocial behaviors/attitudes, rebelliousness, academic performance, perceived risk of drug use, intentions for adulthood substance use, lifetime substance use), *community* (i.e., attachment, prosocial involvement and reward, drug use consequences and antisocial behavior, perceived availability of substances), *school* (i.e., learning environment perceptions, enjoyment, commitment, benefits of learning, truancy), *home* (i.e., parenting practices, family history of substance abuse, rewards for prosocial behavior, parental attitudes regarding antisocial behavior and drug use, relationship quality with parents), and *social* (i.e., best friends engaged in antisocial behavior, tried alcohol or drugs, exhibited prosocial behavior; social rewards for antisocial and prosocial behaviors) influences. See supplementary Table S1 for all items examined in the current study.

#### Outcome

Students reported whether they ever tried electronic cigarettes or e-cigarettes (i.e., *yes* or *no)*. They also reported whether they had ever tried tobacco cigarettes, even just a puff (i.e., *yes* or *no*). Two dichotomous outcome variables were created from these items to represent lifetime exclusive e-cigarette use and dual use (i.e., tobacco and e-cigarette). The comparison group for each outcome were students who did not use either substance.

### Analytic plan

In our sample, 47% (*n* = 6,744) of participants were missing at least one covariate. Prior to imputation, data was randomly resampled into training (70%; *n* = 9,657 e-cigarette; *n* = 9,490 dual) and testing sets (30%; *n* = 4,237 e-cigarette; *n* = 4,065 dual). We then used mode imputation, wherein missing values were replaced with the mode for each variable to address missingness independently for training and testing sets. Mode imputation is commonly utilized within the context of machine learning for classification task [28]. As algorithms can struggle to predict low base rate outcomes, a method known as down sampling was used to randomly resample and reduce the negative class (i.e., those that did not use e-cigarettes or tobacco) until it was equal to the positive class within the training set [27]. Thus, rates of lifetime use and non-use were equal for each outcome within the resampled training sets. The training sets were *n* = 1,108 for exclusive e-cigarette use and *n* = 774 for dual use.

Five dissimilar machine learning algorithms-elastic net, random forest, neural networks, k-nearest neighbors, and logistic regression–were then fitted to the training set to create classification models for each outcome [35]. Each classification algorithm drew information from 112 variables representing student individual and socioecological factors. 5-fold cross-validation was used to identify variables that improved classification accuracy across random subsets of data within the training set [28]. Model performance was assessed on a test set using the Area Under (AUC) of the Receiving Operator Characteristic (ROC) curve, which represents the ability of a model to classify outcomes across all possible cut points [27]. The top performing classification algorithm on the test set was selected for each outcome (i.e., AUC; sensitivity, specificity; [27]). Variable importance figures reflect results from the best performing algorithms for each outcome. High value classifiers were then identified through visual inspection of the relative importance figures. Variables that demonstrate large increase in relative importance over subsequent covariates were said to be high value classifiers [28]. High value classifiers were examined using a crosstabulation visualization to determine the nature of the relationship between each variable and the corresponding outcome [13].

## Results

Patterns of lifetime e-cigarette use differed by demographic variables within the current sample. Chi-square test of independence suggest e-cigarette usage was significantly (*p* < .001) associated with student gender, grade, and race/ethnicity. Boys, 8^th^ graders, Latinxs, Native Americans, and mixed-race students reported the greatest proportion of use across outcomes. Exclusive and dual use were generally associated with a greater proportion of lifetime use across substances relative to no-users. See Table 1 for demographic variables by outcomes.

### Exclusive use

Algorithmic performance on the exclusive e-cigarette use classification task ranged from good to outstanding (AUC = . 787 - .926) on the test set. See supplemental Fig S1 for ROCs for classification algorithms. Elastic net was the best performing algorithm in classifying exclusive e-cigarette use (AUC = .926, sensitivity = .857, specificity = .848). In contrast, logistic regression was the worst performing algorithm in classifying lifetime e-cigarette use (AUC = .787, sensitivity = .768, specificity = .806). Elastic net identified perceived availability of e-cigarettes, lifetime alcohol use, parents’ attitudes regarding their use of vape products, school suspension, perceived risk of e-cigarette use, lifetime marijuana use, best friend(s) tried alcohol, best friend(s) used marijuana, and perceived risk of smoking marijuana regularly as the best discriminators between lifetime exclusive e-cigarette users and non-users. See Fig 1 for variable importance. Visual inspection of cross-tabulation mosaics suggests that perceived availability of e-cigarettes (i.e., *sort of hard*, v*ery easy, sort of easy*), lifetime substance use (i.e., alcohol, marijuana), school suspensions (i.e., 1 or more), lower levels of perceived risk associated with e-cigarette use (i.e., *none* to *moderate*), best friend(s) tried alcohol or used marijuana (i.e., 1 or more), and less perceived risk associated with smoking marijuana regularly were all associated with a greater proportion of lifetime e-cigarette use. Students who reported that their parents would view their use of vape products as “*very wrong*” had the lowest proportion of use relative to other levels of approval (i.e., *wrong* to *not wrong at all*). See supplemental Fig S3-11 for cross-tabulation visualizations.

**Fig 1.**
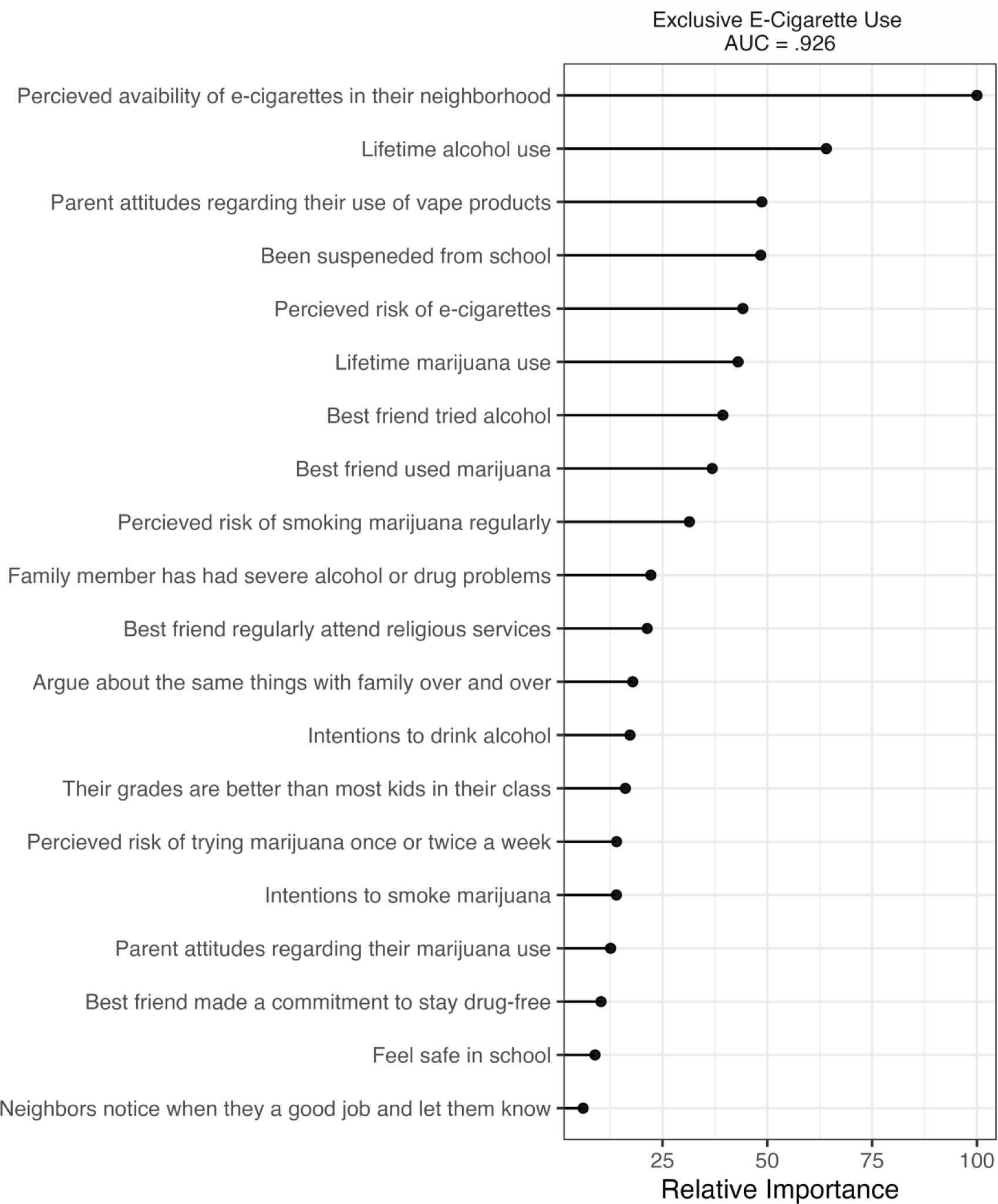
Top 20 Variables with the Highest Relative Importance in Classifying Lifetime E-Cigarette Use. Results represent validation on a separate test dataset.

### Dual use

Algorithmic performance on the dual tobacco and e-cigarette use classification task also ranged from excellent to outstanding (AUC = . 725 - .944) on the test set. See supplemental Fig S2 for ROCs for classification algorithms. Elastic net and random forest had the same AUC score (.944). However, elastic net (sensitivity = .824, specificity = .947) outperformed random forest (sensitivity = .818, specificity = .939) based on sensitivity and specificity. Logistic regression was the worst performing algorithm in classifying lifetime dual use (AUC = .725, sensitivity = .630, specificity = .779). Elastic net identified lifetime alcohol use, lifetime marijuana use, perceived availability of e-cigarettes, best friend(s) cigarette use, perceived risk of e-cigarette use, lifetime inhalants use, school suspension, and perceived risk of smoking marijuana regularly as the best discriminators between lifetime dual users and non-users. See Fig 2 for variable importance. Visual inspection of cross-tabulation mosaics suggests that lifetime substance use (i.e., alcohol, marijuana, inhalants), higher levels of perceived availability of e-cigarettes (i.e., v*ery easy, sort of easy*), best friend(s) that have smoked cigarettes (i.e., 1 or more), school suspensions (i.e., 1 or more), lower levels of perceived risk associated with e-cigarette use and using marijuana regularly (i.e., *none* to *moderate*) were all associated with a greater proportion of lifetime dual use. See supplemental Fig S12-19 for cross-tabulation visualizations.

**Fig 2.**
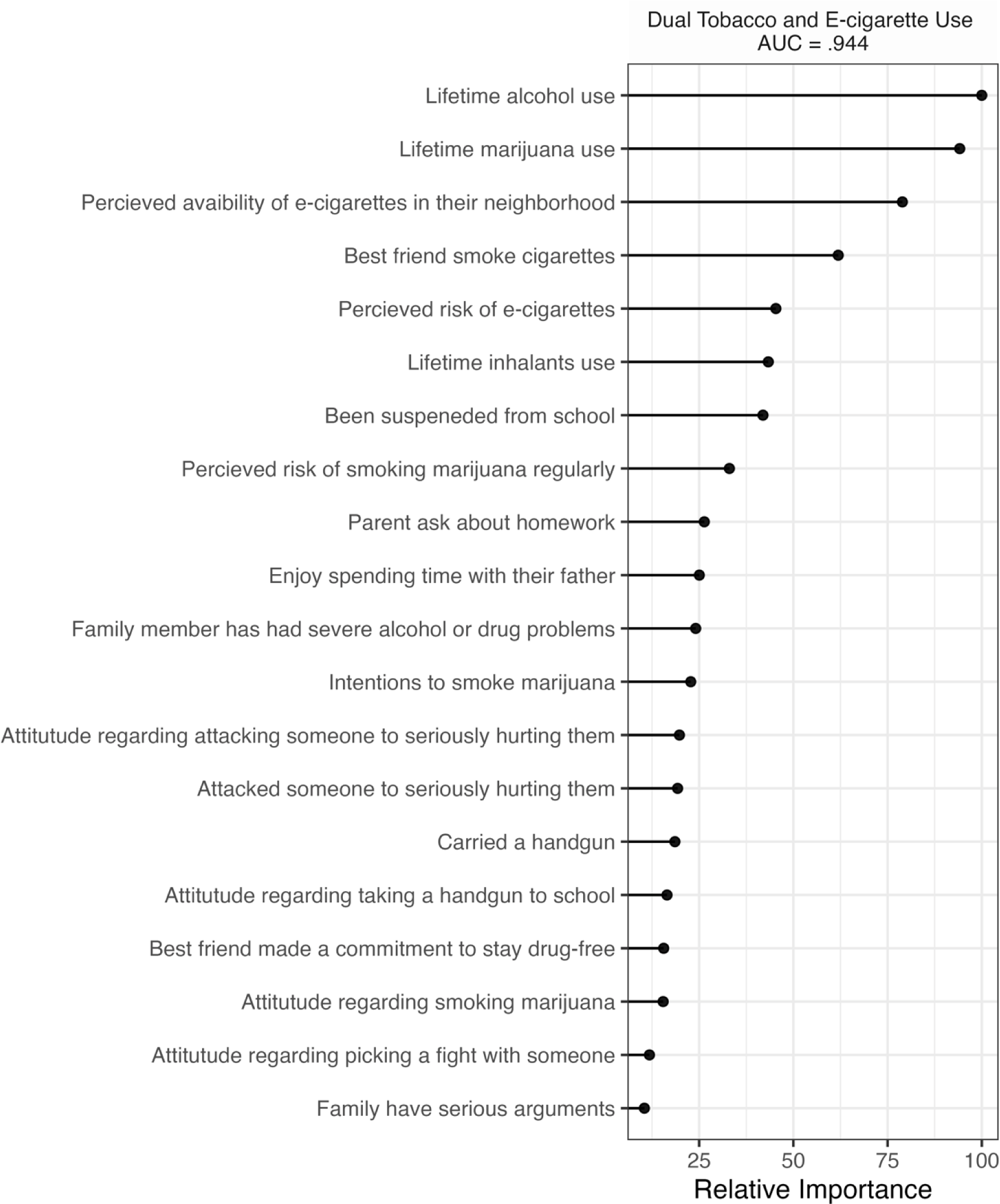
Top 20 Variables with the Highest Relative Importance in Classifying Lifetime Dual Use. Results represent validation on a separate test dataset.

## Discussion

The current study expands the literature through the simultaneous exploration of established correlates of e-cigarette initiation and traditional factors associated with substance use in relation to early adolescent vaping. Algorithms utilizing information regarding student individual characteristics and socioecological context demonstrated high levels of classification accuracy for both lifetime exclusive and dual e-cigarette use. Elastic net generally outperformed other algorithm in classification accuracy. While the order of importance of classifiers differed by outcome, elastic net consistently identified six high value classifiers across usage groups: lifetime alcohol or marijuana use, perception of e-cigarette availability and risk, school suspension(s), and perceived risk of smoking marijuana regularly. Several high value classifiers differed between youth who reported lifetime exclusive (i.e., parent’s attitudes regarding their use of vaping products, best friend[s] tried alcohol, best friend[s] used marijuana) and dual e-cigarette use (i.e., best friend[s] smoked cigarettes, lifetime inhalants use). These findings highlight important commonalities and difference in risk profiles between lifetime exclusive and dual e-cigarette users.

Research using high school samples have documented higher rates of life substance use among dual versus exclusive e-cigarette users [7]. Within our sample, rates of substance use were generally higher among exclusive and dual users relative to those who abstained from both. Consistent with prior research, the greatest portions of substance use were found among youth who had reported dual use [26]. However, only lifetime alcohol and tobacco were found to be important classifiers of both e-cigarette use outcomes among middle school students, which is consistent with prior findings in high school samples [7]. Our findings also extend prior work through the identification of inhalant use as a novel risk factor specifically related to lifetime dual use during early adolescence. It is possible that dual users may access a wide variety of substances recreationally and may utilize inhalants as they are easy to access within the home [26,36]. Further research is needed to understand the relationship between lifetime inhalant and dual use.

Our findings confirm that availability of e-cigarettes is an important influence on early adolescent vaping [19], despite a Utah state law that restrict the sale of these products to individuals under the age of 20. Accessibility was especially important to exclusive e-cigarette use, which is concerning as this may translate to future traditional cigarette use among adolescents who may have otherwise abstained from tobacco use [8]. Consistent with prior research, students who reported lower perceived danger of using e-cigarettes reported a greater proportion of lifetime use [12]. Our findings suggest a need to consider perceptions regarding the danger of other substances, such as marijuana, when assessing risk for early adolescent e-cigarette use. While recent findings have highlighted the importance of school-based factors in assessing risk for e-cigarette using among high school samples (i.e., truancy and poor academic performance; [10]), our findings suggest that student suspensions were the most relevant aspect of school in relation to early adolescent e-cigarette use. It is possible school suspensions may be associated with an increased risk for e-cigarette use as a potential proxy for rule breaking behaviors or through greater unsupervised time outside of school [7,37]. Further research is needed to elucidate the relationship between school suspensions and early adolescent e-cigarettes use.

It is important to mention that factors traditionally associated with substance use such as adolescent and peer delinquency, community substance use norms, and school involvement were not important predictors of lifetime exclusive and dual e-cigarette use within the current sample. These findings may signal potential differences between factors underlying e-cigarette and other forms of substance use. Additionally, factors identified by prior research as relevant predictors of vaping (e.g., parenting practices, perceived risk of smoking tobacco) were not relevant correlates of patterns of e-cigarette use within the current sample [7,12]. It is possible that when competing against other variables within a machine learning approach, these important predictors are truly of lesser importance relative to high value classifiers identified in the current study.

### Implications

Our findings support addressing early adolescent vaping through prevention programs aiming to address substance use prevention broadly (i.e., alcohol, tobacco, marijuana, inhalants, e-cigarette). There is ample evidence for the efficacy of prevention programs whether focused on a single substance or multiple ones [38]. Our findings provide specific structural and behavioral targets that may inform the adaptation of programs seeking to prevent different patterns of early adolescent e-cigarette use. Substance use prevention programs may benefit from adding components that equip parents to effectively communicate disapproval regarding their child’s use of e-cigarette and teach youth skills to resist peer substance use influences [39,40]. Targeted prevention programs are already supported by research documenting perception of risk associated with vaping as a consistent predictor of e-cigarette use [7,8] and our findings highlight the importance of also considering perceptions of danger regarding marijuana use during early adolescence. Specific to programming, youth’s perceptions of risk do not align with research evidence providing an important point of content for preventive interventions [41]. Altering youth’s perception of e-cigarette accessibility, however, may require intervention at a broader social level (e.g., public media campaigns). Alternatively, decreasing accessibility to e-cigarettes may be achieved by actions external to youths such as strong enforcement of laws regarding possession and/or consumption for underage users and/or those selling e-cigarette products to them, or by way of increasing prices for goods associated with e-cigarette use.

Machine learning appears to be a promising screening tool for the identification of risk factors that can accelerate the development of the e-cigarette knowledge base needed to curb the rapid spread of vaping among adolescents nationally. Algorithms were able to efficiently explore a wide range of factors in association with early adolescent e-cigarette use, which confirmed findings from later developmental stages and identified several novel risk factors (i.e., inhalant use, perceived risk of marijuana, school suspensions). An important consideration in this research is that the tools utilized to identify e-cigarette use classifiers are publicly available. R offers open access statistical packages for machine learning. Additionally, there are substantial training materials available for free online. These tools can provide an accessible and replicable method of generating and disseminating scientific knowledge regarding e-cigarette use classifiers nationally. We encourage researchers to apply machine learning algorithms to their data to draw new insight regarding factors contributing to a variety of e-cigarette use outcomes among adolescents. Examining cross-sectional markers of e-cigarette use could also identify important variables that can be examined longitudinally in prospective research. Machine learning algorithms have many exciting applications when applied to longitudinal data, including identifying context specific predictors of service use, specific targets for substance use prevention programs, and ensure that important factors are not excluded from causal models examining mechanism underpinning early vaping initiation.

### Limitations

Results from algorithms used in the current study do not necessarily imply causal mechanisms explaining patterns of lifetime e-cigarette use but rather identify factors that are strong correlates of group membership (i.e., use or non-use). Longitudinal research is needed to establish causal links. The current study examined lifetime substance use that may range from experimentation to habitual use. Future research may consider using machine learning as a method of identifying youth at-risk for habitual e-cigarette use. Although a large number of the Utah adolescent population was captured, the PNA survey does not include students in private schools, correctional facilities, or treatment centers. Additionally, students who were not in attendance, declined participation, or did not return parental consent forms are not represented. Furthermore, findings may not generalize to students in other states. Further research is needed to replicate our findings in different contexts and developmental periods.

### Conclusions

The current study utilized a machine learning approach to efficiently explore and identify high value correlates of early adolescent lifetime e-cigarette use. This approach identified several shared risk factors for exclusive and dual e-cigarette use such as lifetime use of specific substances (i.e., alcohol, marijuana), perception of e-cigarette availability and risk, school suspension(s), and perceived risk of smoking marijuana regularly. Several differences were also identified between youth who reported lifetime exclusive (i.e., parent’s attitudes regarding their use of vaping products, best friend[s] tried alcohol or used marijuana) and dual use (i.e., best friend[s] smoked cigarettes, lifetime inhalants use) relative to non-users. This information provides a first step towards identifying youth at-risk for e-cigarette use during early adolescence. Further research is needed to examine high value classifiers identified by the current study using explanatory models and longitudinal data to understand mechanism underlying their importance in accounting for differences in risk profiles between e-cigarette usage groups during early adolescence.

## Reporting

### Funding

This research was funded by a collaboration between multiple state agencies in Utah (i.e., Department of Health, Department of Human Services, and the State Board of Education).

### Disclosure Statement

The authors have no conflicts of interest associated with the publication of this manuscript.

## Data Availability

We are not authorized to share the Utah Prevention Needs Assessment data used in the current study as it is owed and managed by the Utah Department of Human and Health Services. Data used in the current study can be requested from the Utah Department of Human and Health Services.

## Data Availability

Data used in the current study can be requested from the Utah Department of Human and Health Services.

## Data Deposition

We are not authorized to share the Utah Prevention Needs Assessment data used in the current study as it is owed and managed by the Utah Department of Human and Health Services.

## Notes

### Competing Interest Statement

The authors have declared no competing interest.

### Author Declarations

The Utah State University Institutional Review Board approved secondary analyses of the 2017 Utah PNA survey data as non-human subjects research as participants could not be re-identified (protocol #10108).

### Summary of Updates

Minor corrections to typographical errors.

