## Supplemental for "An ecological examination of early adolescent e-cigarette use: A machine learning approach to understanding a health epidemic"

**Fig S1. Receiver Operating Characteristics (ROC) Curve Estimating Area Under the Curve (AUC) for Models Classifying Exclusive Lifetime E-cigarette Use.** EN = Elastic Net, KNN = K-nearest neighbors, NN = Neural networks, RF = Random forest, LR = Logistic regression.

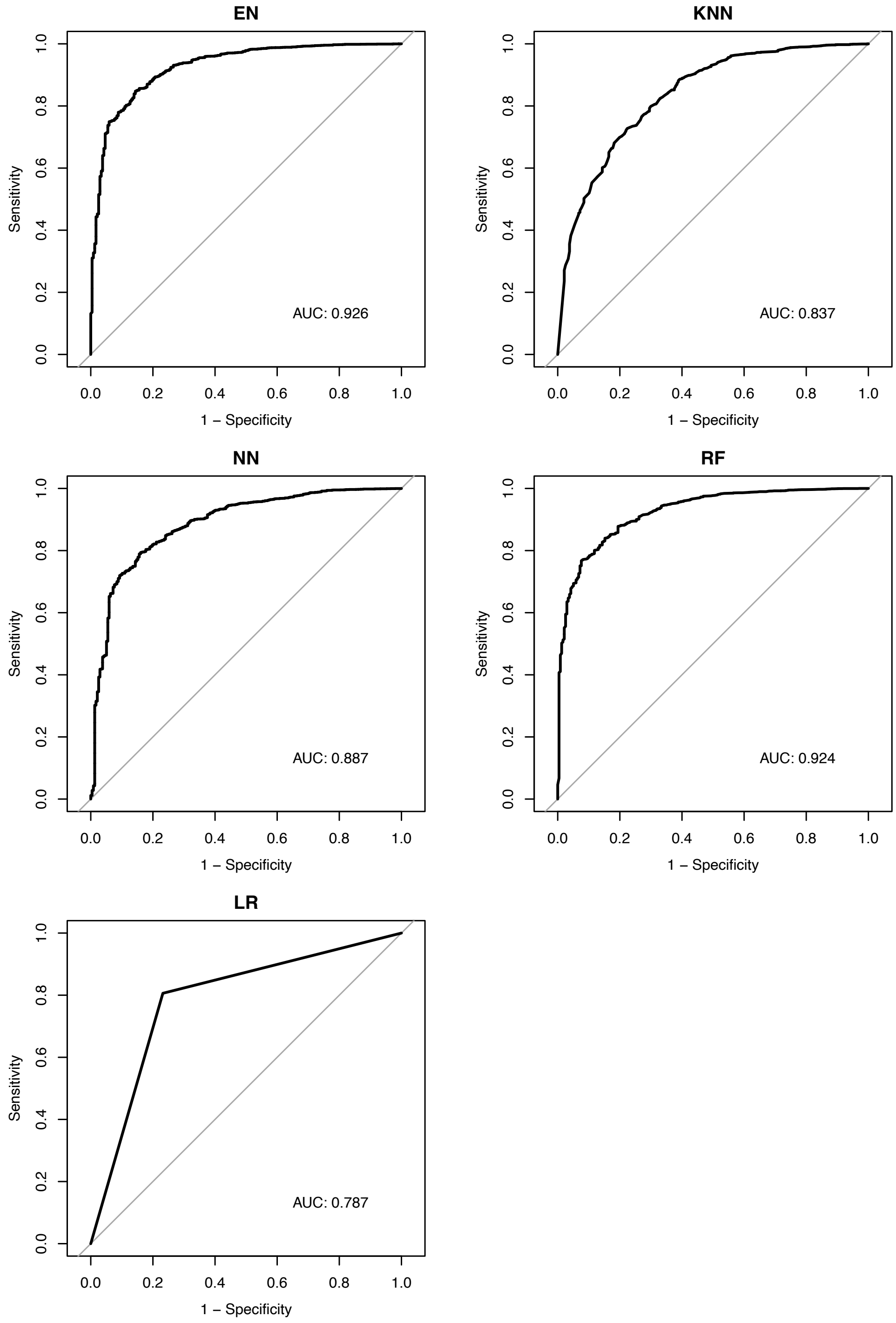

**Fig S2. Receiver Operating Characteristics (ROC) Curve Estimating Area Under the Curve (AUC) for Models Classifying Dual Lifetime Tobacco and E-cigarette Use.** EN = Elastic Net, KNN = K-nearest neighbors, NN = Neural networks, RF = Random forest, LR = Logistic regression.

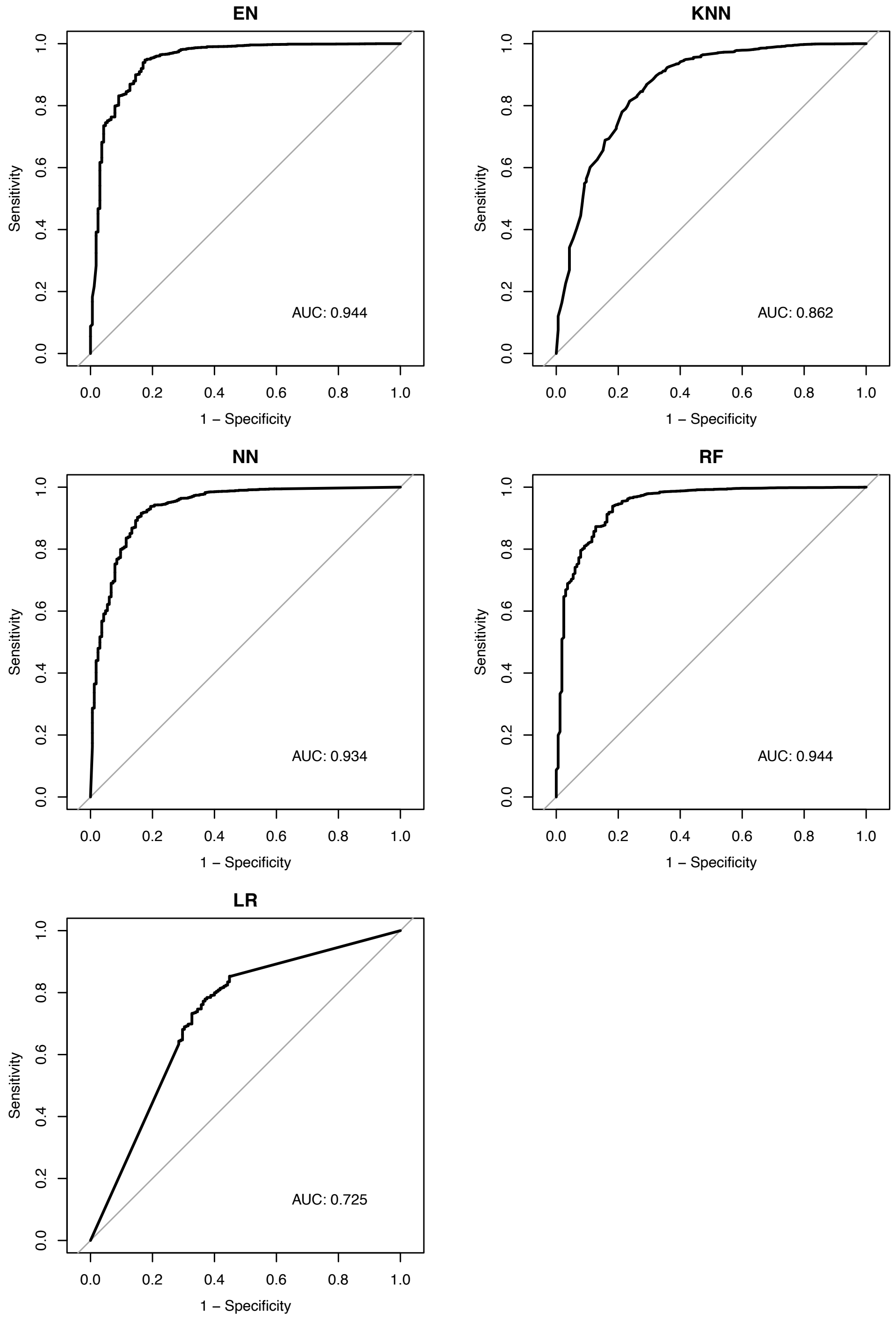

**Fig S3. Cross-Tabulation Visualization Showing Proportion of Exclusive Lifetime E-cigarette Use by Perceived Availability of E-cigarettes.** *VH = Very hard, SH = Sort of Hard, SE = Sort of easy, VE = Very easy*

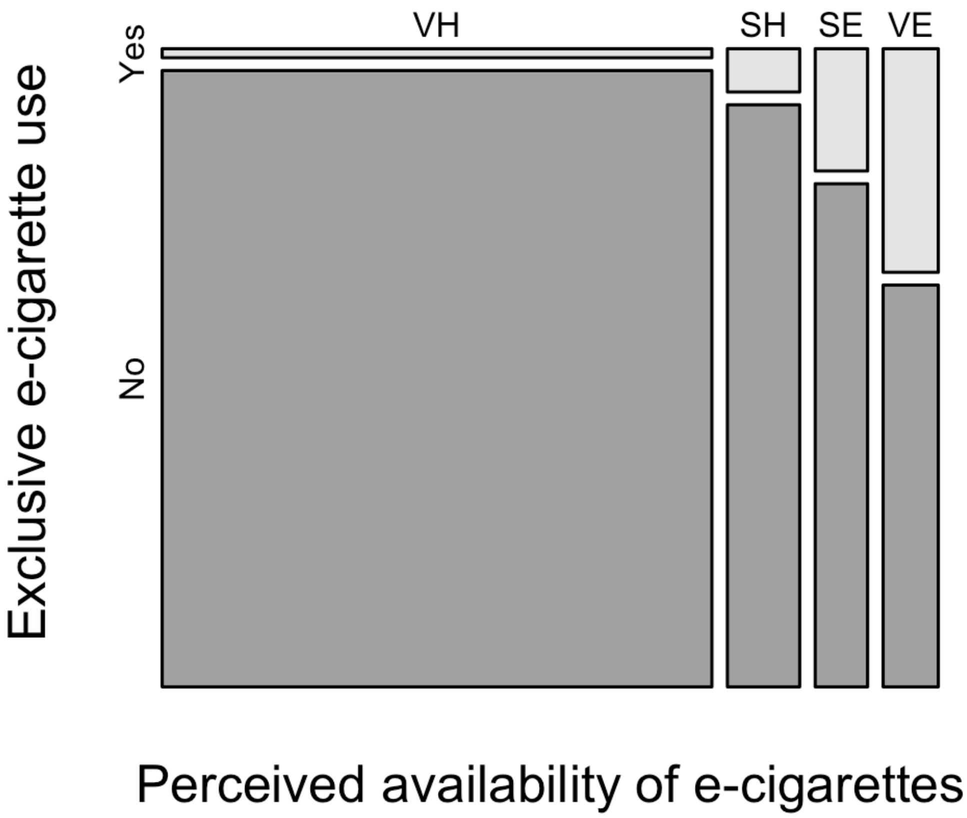

**Fig S4. Cross-Tabulation Visualization Showing Proportion of Exclusive Lifetime E-cigarette Use by Lifetime Alcohol Use.**

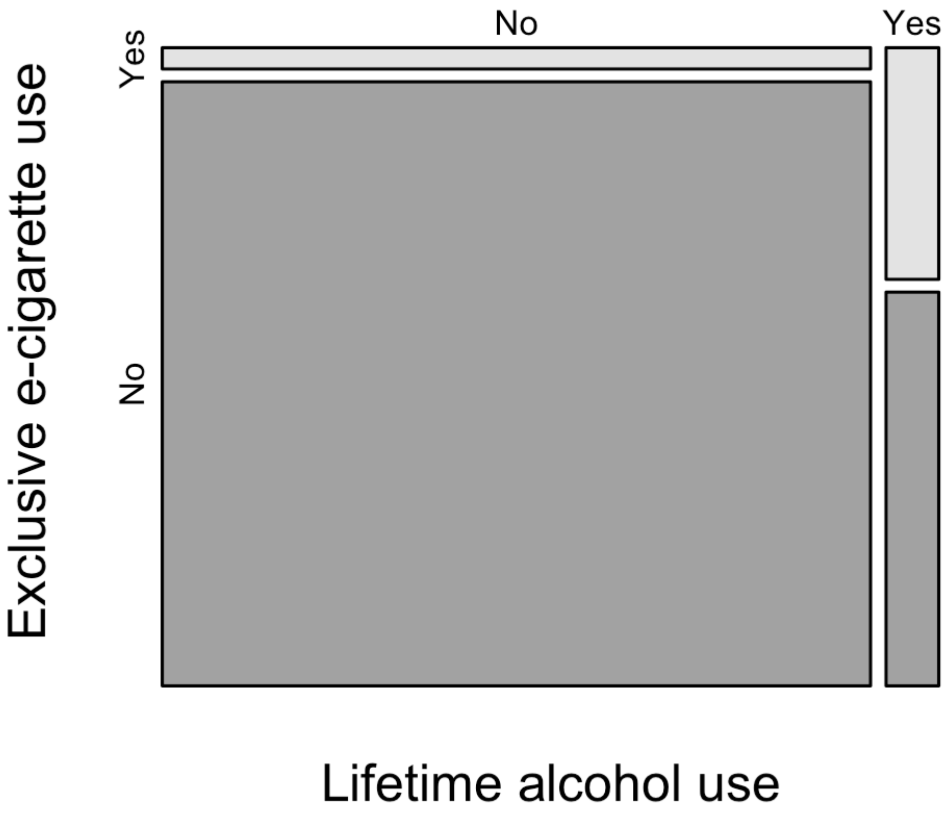

**Fig S5. Cross-Tabulation Visualization Showing Proportion of Exclusive Lifetime E-cigarette Use by Parents Attitudes Regarding their Use of Vape Products.** V = Very wrong, W = Wrong, A = A little bit wrong, N = Not wrong at all.

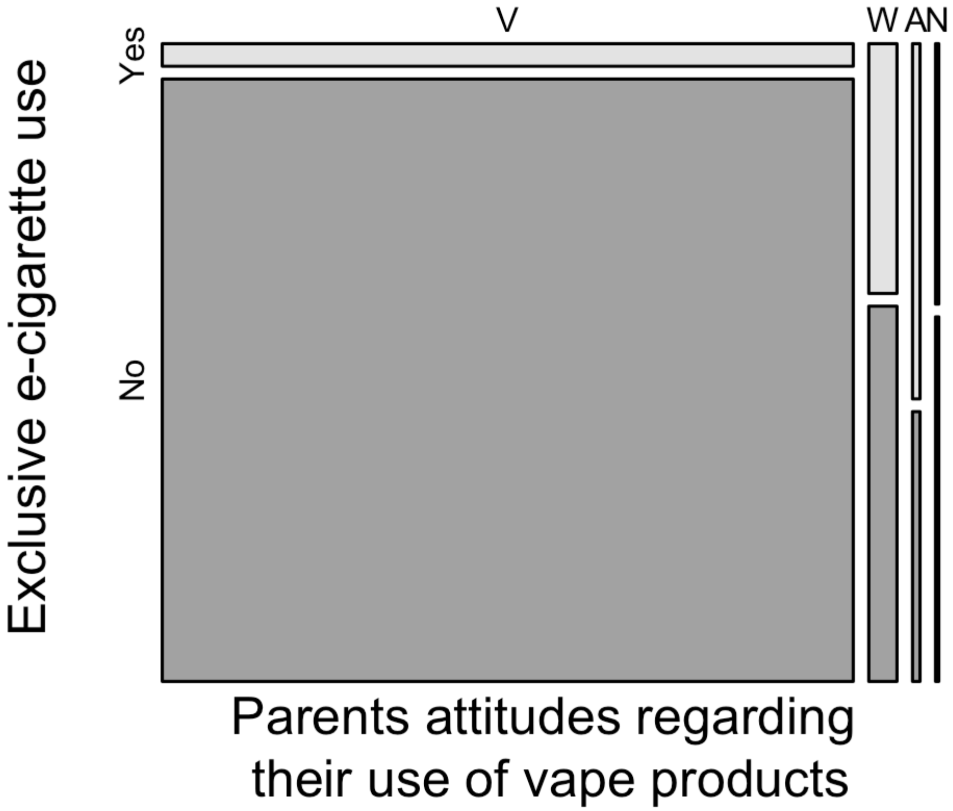

**Fig S6. Cross-Tabulation Visualization Showing Proportion of Exclusive Lifetime E-cigarette Use by School Suspension.** 1-2 = 1 or 2 times, + = 3 or more times.

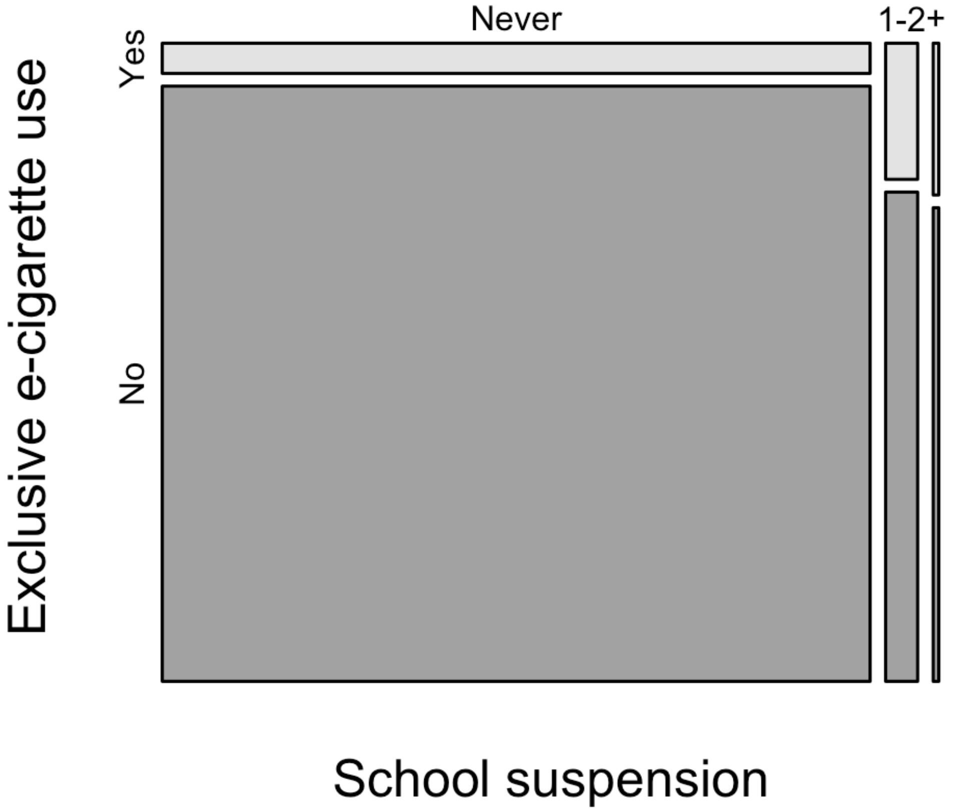

**Fig S7. Cross-Tabulation Visualization Showing Proportion of Exclusive Lifetime E-cigarette Use by Perceived Risk of E-cigarettes.** G = Great Risk, M = Moderate Risk, S = Slight Risk, N = No Risk.

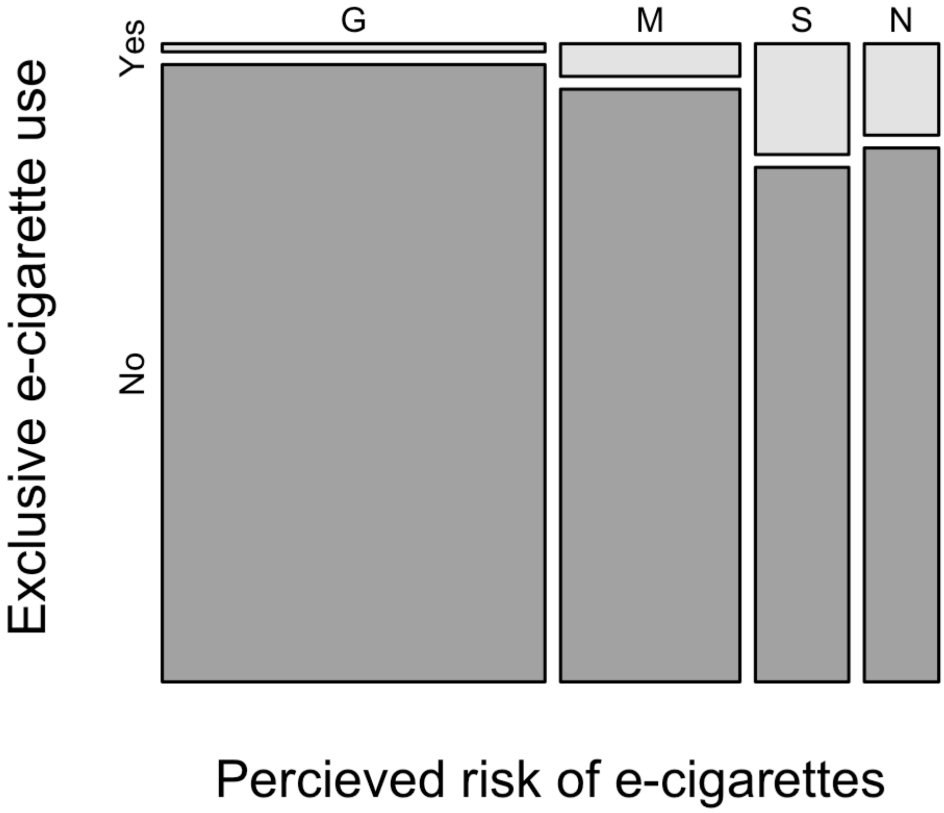

**Fig S8. Cross-Tabulation Visualization Showing Proportion of Exclusive Lifetime E-cigarette Use by Lifetime Marijuana Use.**

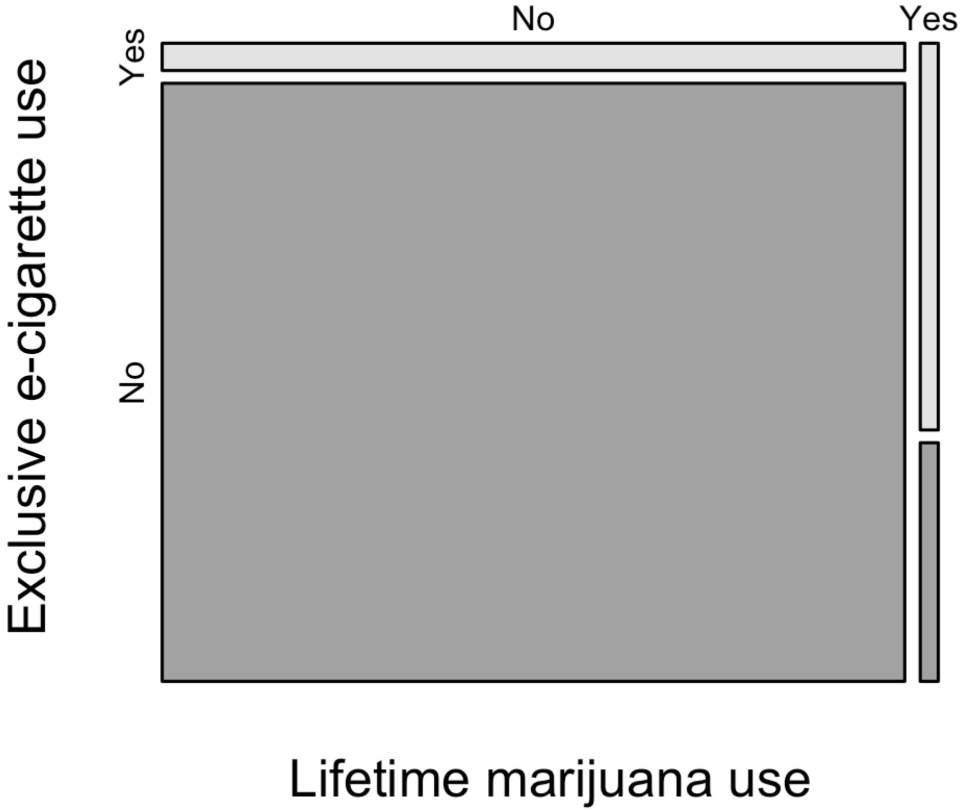

**Fig S9. Cross-Tabulation Visualization Showing Proportion of Exclusive Lifetime E-cigarette Use by Best Friend(s) tried alcohol.** *1-2 = 1 or 2 friends, 3-4 = 3 or 4 friends.*

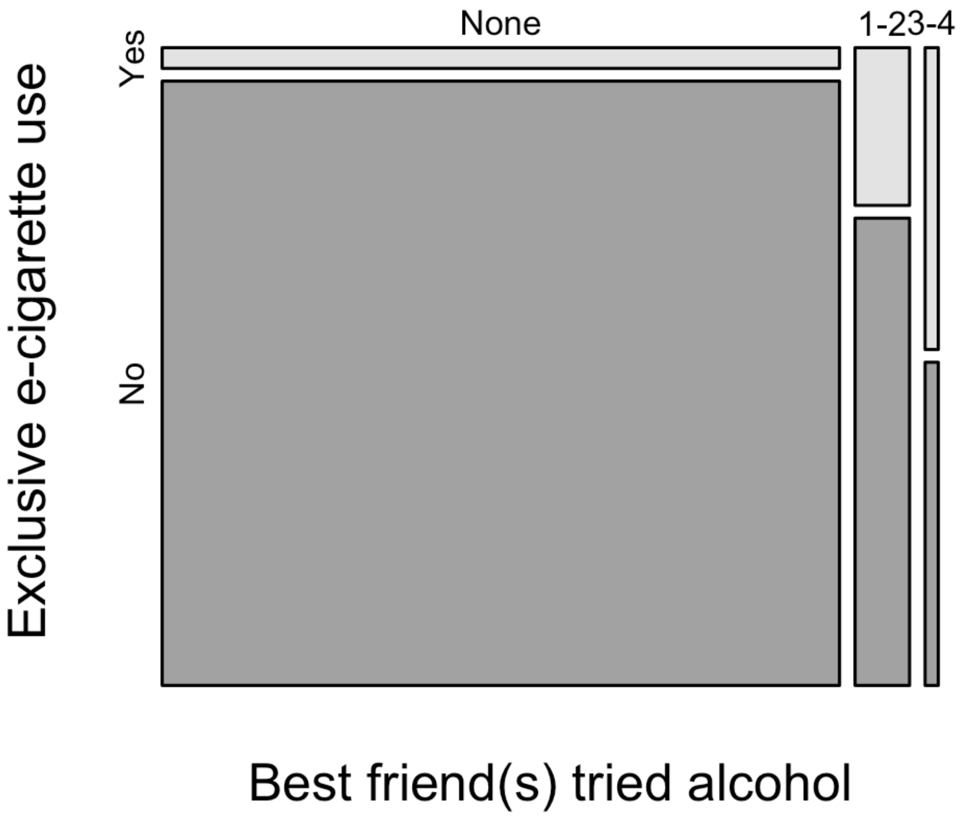

**Fig S10. Cross-Tabulation Visualization Showing Proportion of Exclusive Lifetime E-cigarette Use by Best Friend(s) Used Marijuana.** 1-2 = 1 or 2 friends, 3-4 = 3 or 4 friends.

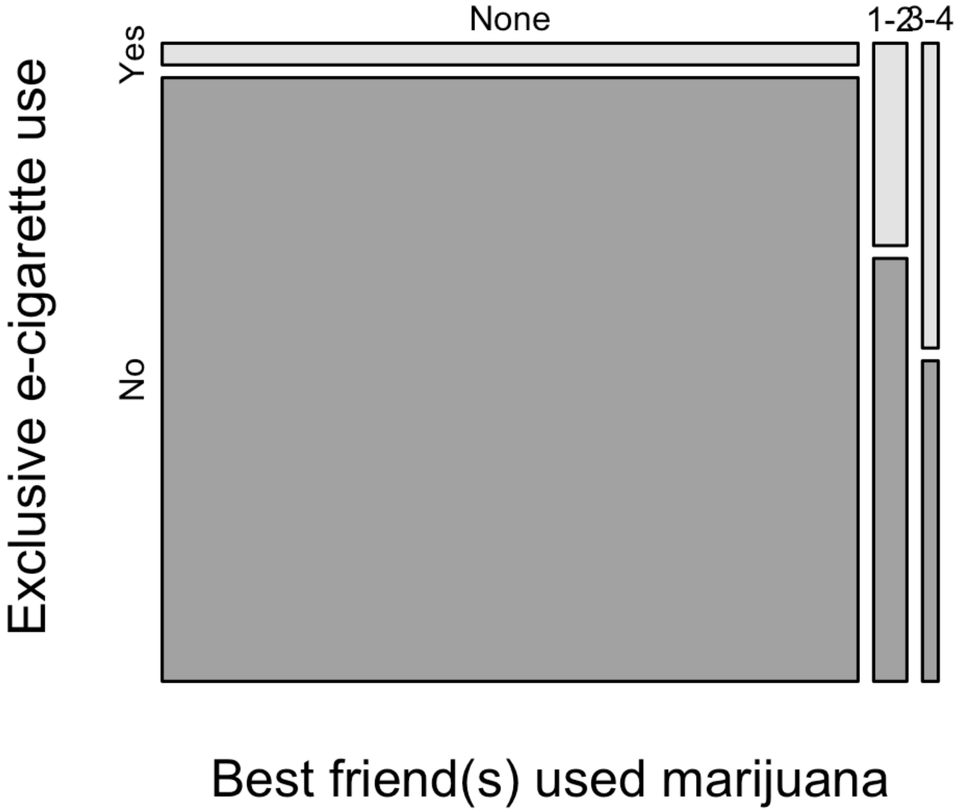

**Fig S11. Cross-Tabulation Visualization Showing Proportion of Exclusive Lifetime E-cigarette Use by Perceived Risk of Smoking Marijuana Regularly.** G = Great Risk, M = Moderate Risk, S = Slight Risk, N = No Risk.

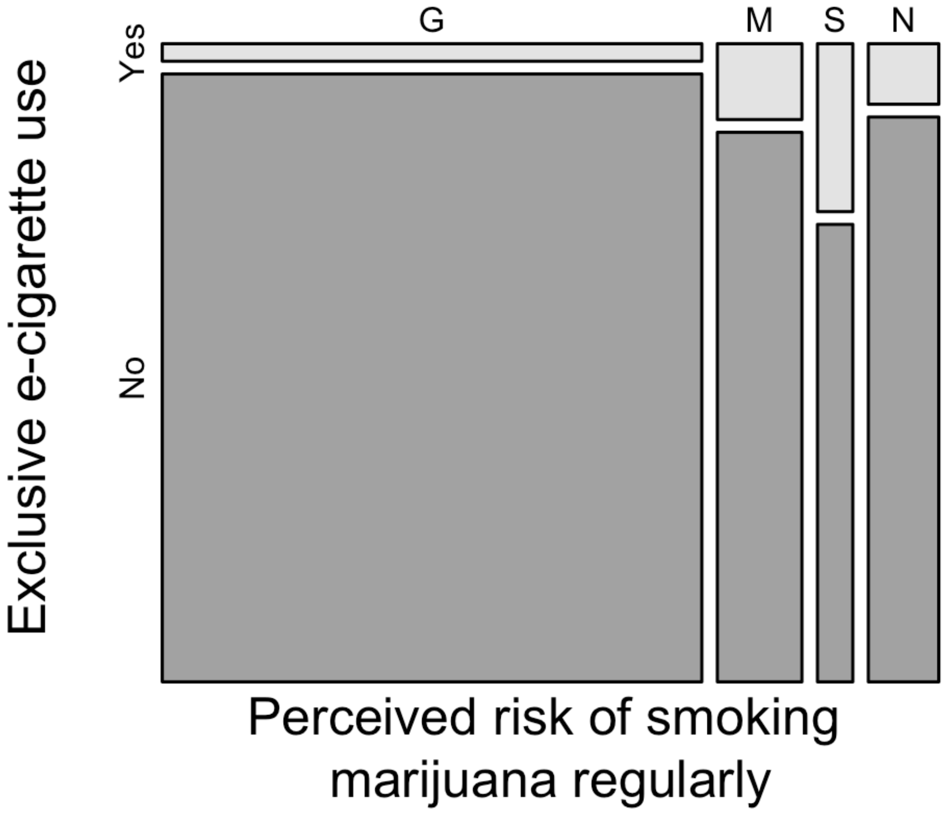

**Fig S12. Cross-Tabulation Visualization Showing Proportion of Dual Lifetime Tobacco and E-cigarette Use by Lifetime Alcohol Use.**

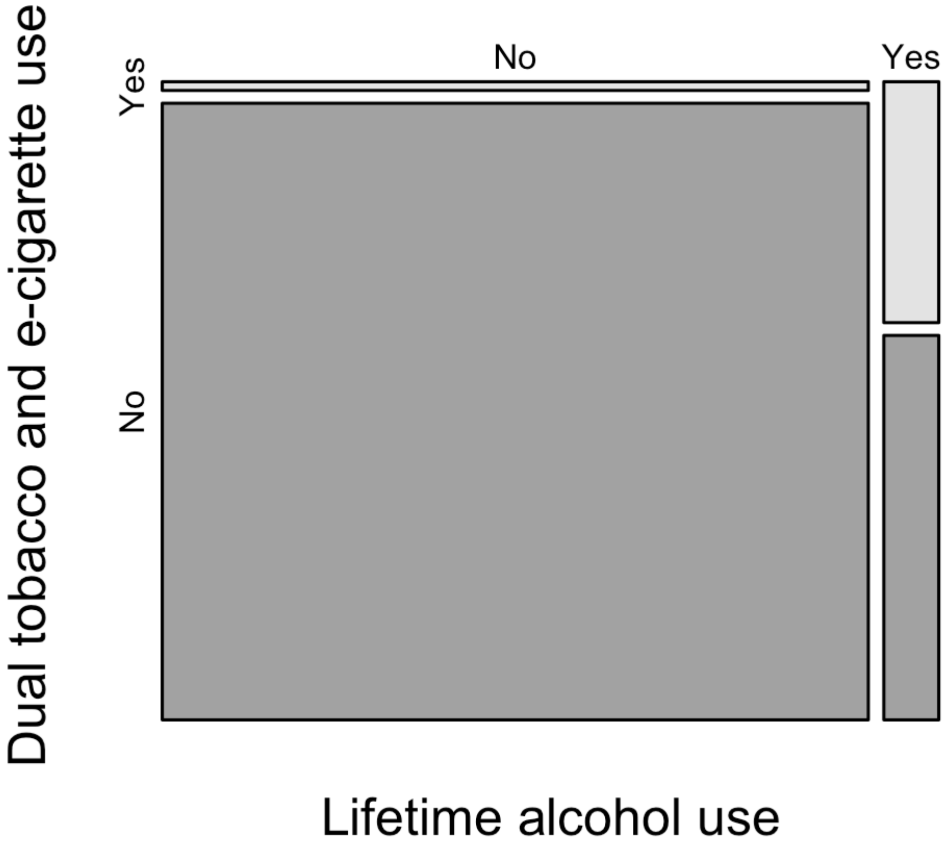

**Fig S13. Cross-Tabulation Visualization Showing Proportion of Dual Lifetime Tobacco and E-cigarette Use by Lifetime Marijuana Use.**

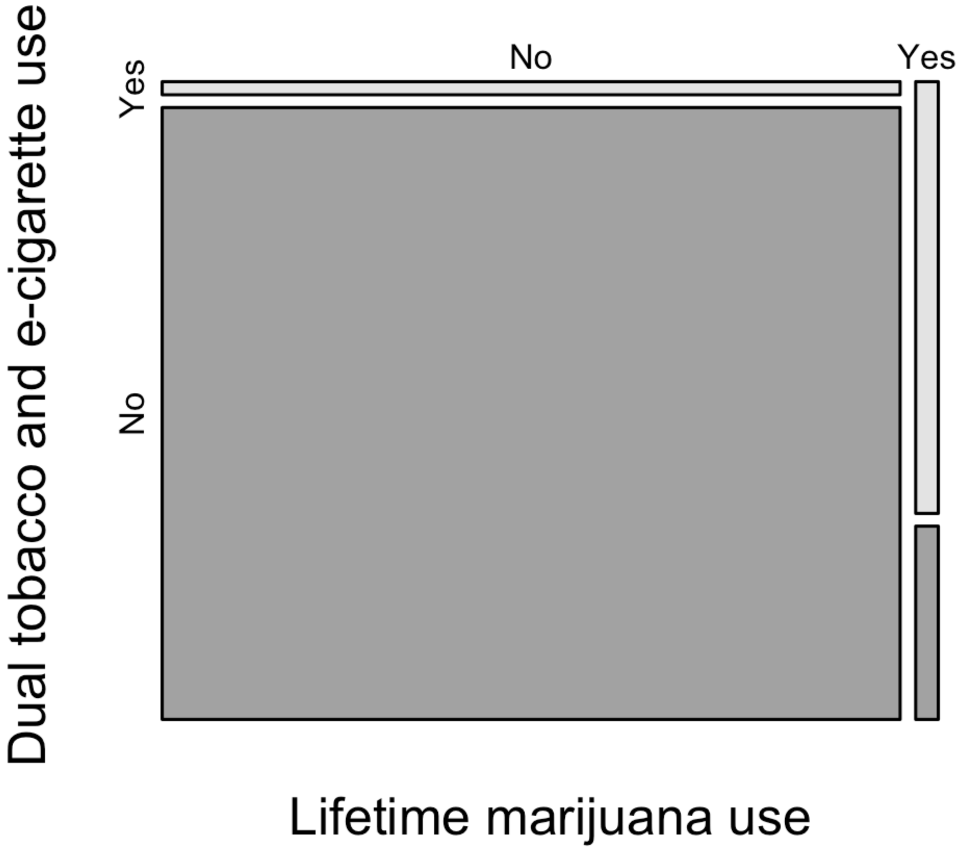

**Fig S14. Cross-Tabulation Visualization Showing Proportion of Dual Lifetime Tobacco and E-cigarette Use by Perceived Availability of E-cigarettes.** VH = Very hard, SH = Sort of Hard, SE = Sort of easy, VE = Very easy.

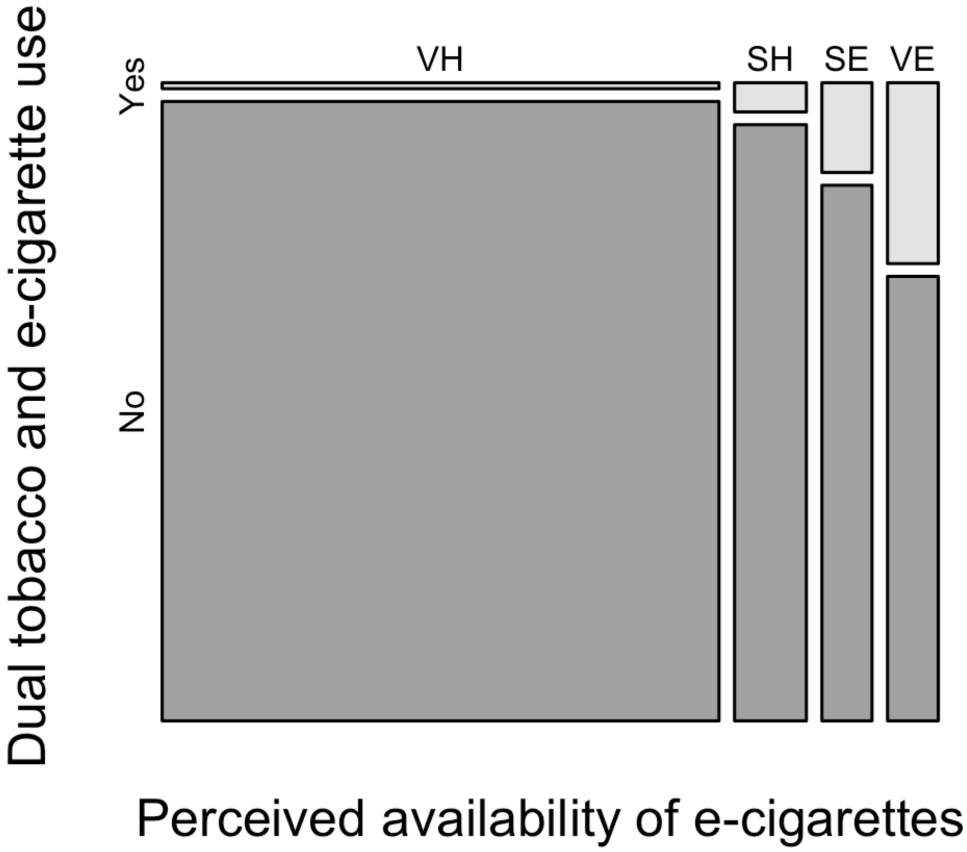

**Fig S15. Cross-Tabulation Visualization Showing Proportion of Dual Lifetime Tobacco and E-cigarette Use by Best Friend(s) Smoked Cigarettes.** 1-2 = 1 or 2 friends, 3-4 = 3 or 4 friends.

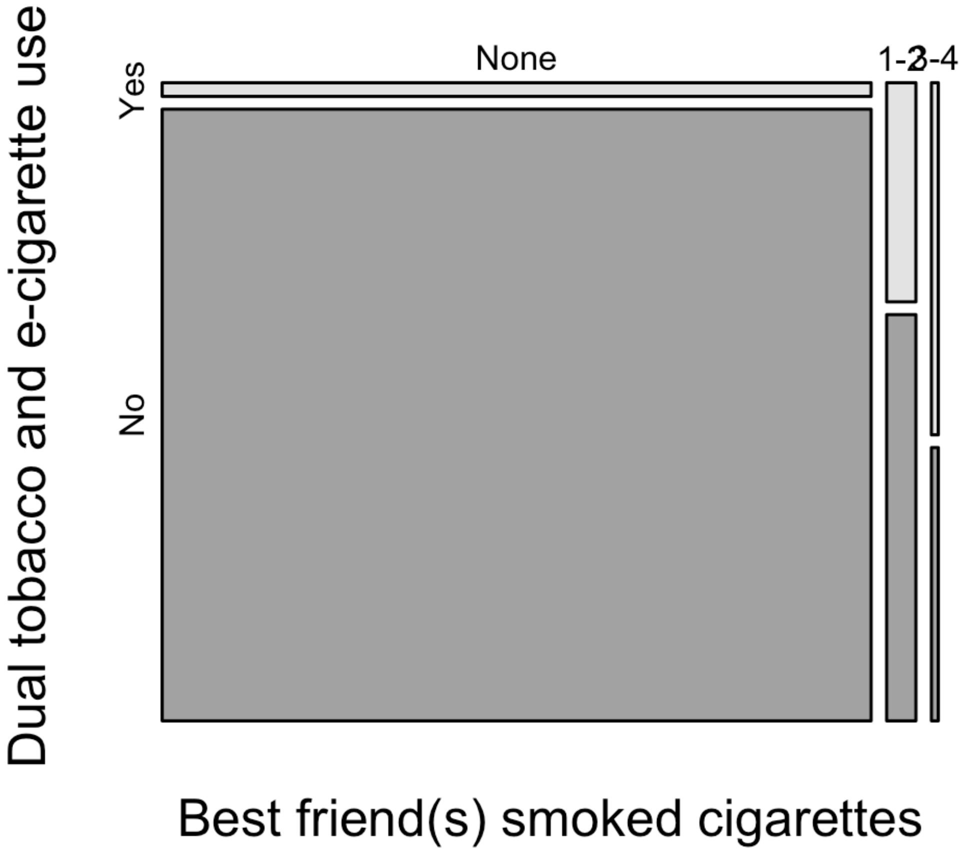

**Fig S16. Cross-Tabulation Visualization Showing Proportion of Dual Lifetime Tobacco and E-cigarette by Perceived Risk of E-cigarettes.** G = Great Risk, M = Moderate Risk, S = Slight Risk, N = No Risk.

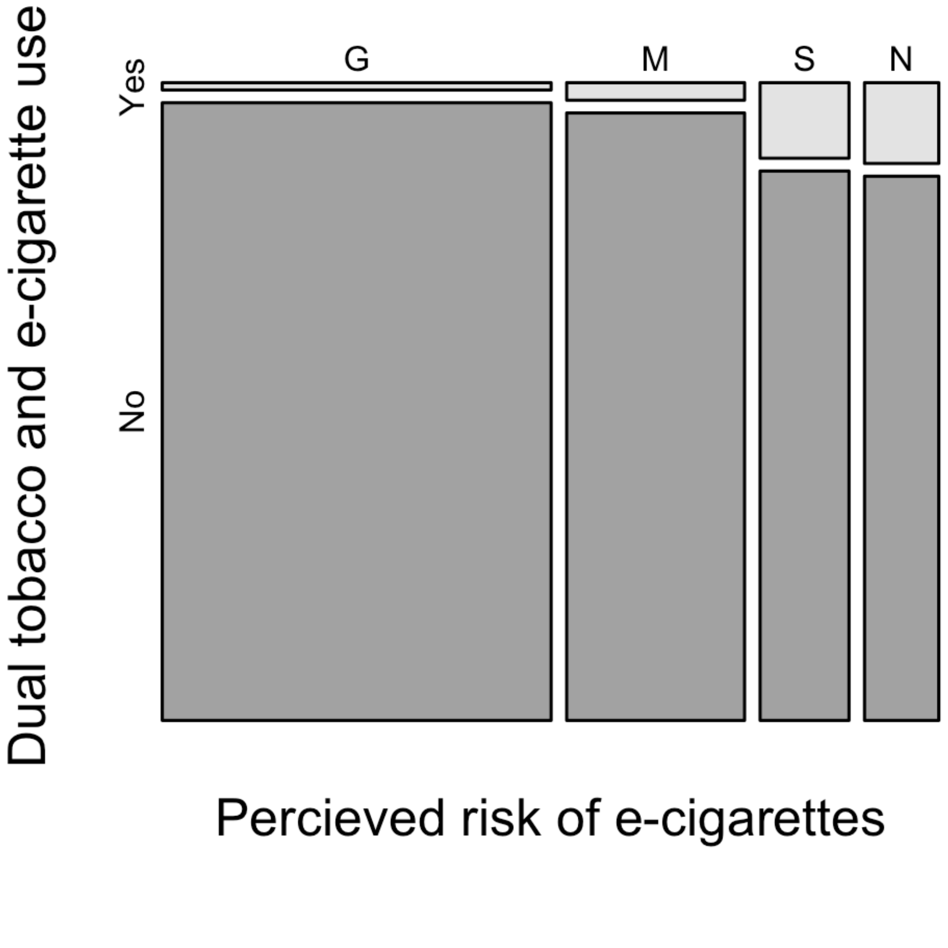

**Fig S17. Cross-Tabulation Visualization Showing Proportion of Dual Lifetime Tobacco and E-cigarette by Lifetime Inhalants Use.**

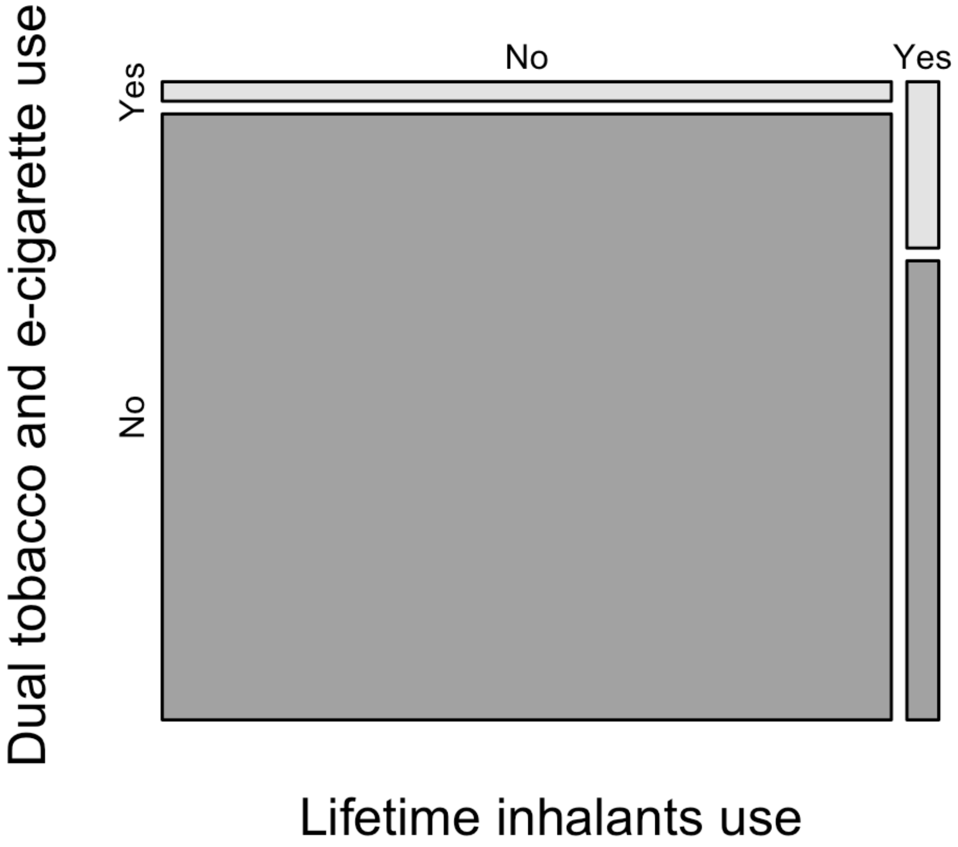

**Fig S18. Cross-Tabulation Visualization Showing Proportion of Dual Lifetime Tobacco and E-cigarette by School Suspension.** 1-2 = 1 or 2 times, + 3 or more times.

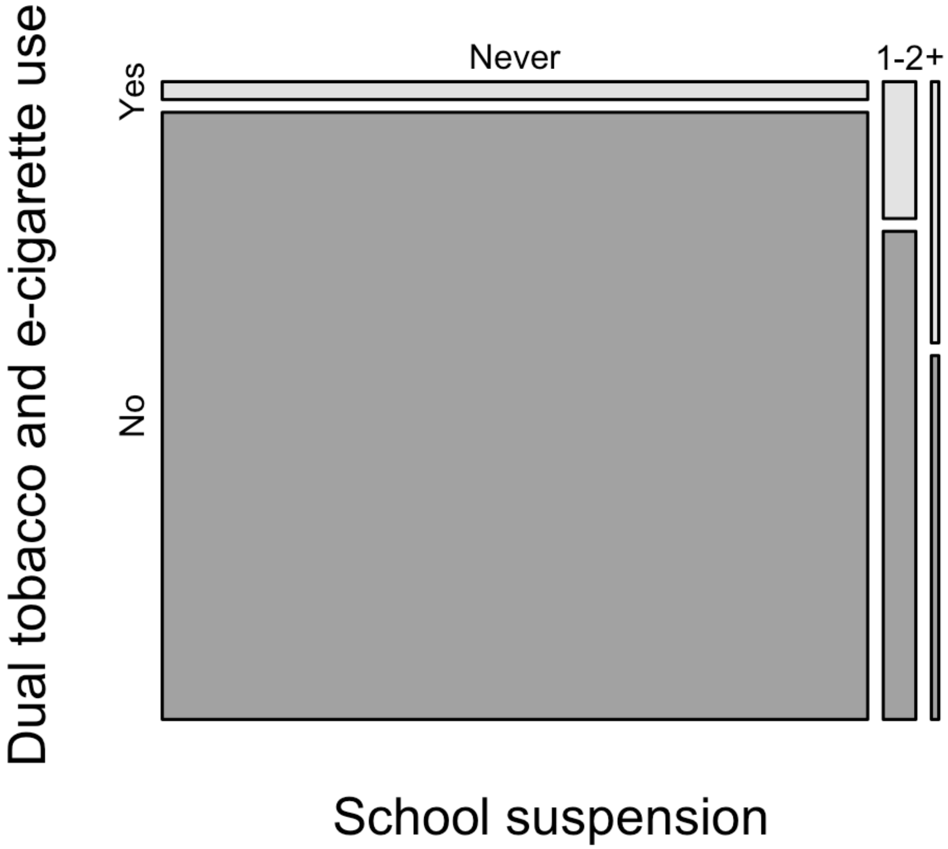

**Fig S19. Cross-Tabulation Visualization Showing Proportion of Dual Lifetime Tobacco and E-cigarette by Perceived Risk of Smoking Marijuana Regularly.** G = Great Risk, M = Moderate Risk, S = Slight Risk, N = No Risk.

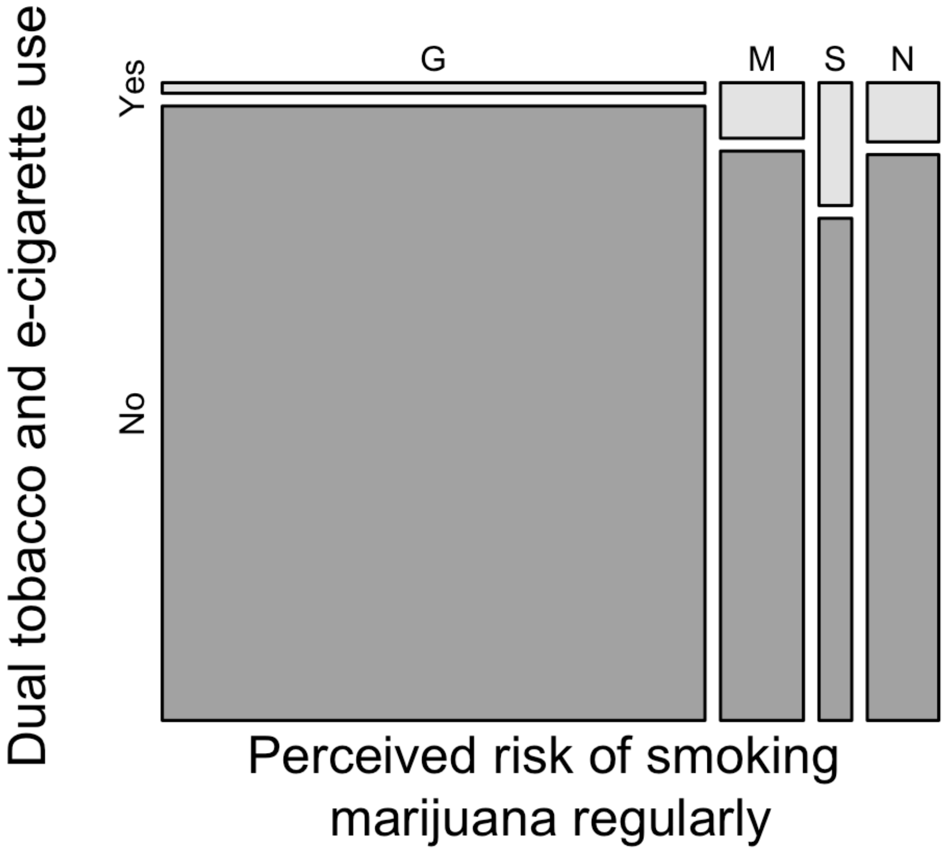
